# Association between Serum Prolactin and Metabolic Disorders: a Systematic Review

**DOI:** 10.1101/2020.08.25.20180182

**Authors:** Zaibo Yang, Junsen She, Congcong Zhou, Mu Liangshan

**Affiliations:** School of Medicine, Zhejiang University, Hangzhou, 310000, P. R. China; First Clinical Medical College, Wenzhou Medical University, Wenzhou, P. R. China

**Keywords:** Prolactin, Type 2 Diabetes Mellitus, Obesity, Hypertension, Insulin Resistance, Systematic Review

## Abstract

**Background:** Serum prolactin levels are associated with metabolic disorders. However, the conclusions were inconsistent among published studies.

**Methods:** PubMed, EMBASE, and the Cochrane Library were used to search for studies investigating the association between serum prolactin levels and metabolic disorders. Studies were included and reviewed if they reported the association between serum prolactin and metabolic components (including waist circumference, body weight indexes, blood pressure, blood glucose, blood lipids, insulin resistance, and type 2 diabetes).

**Results:** A total of 14 studies were included in this systematic review. Evidence for certain associations between serum prolactin levels and body weight, blood lipids, blood glucose was insufficient, while some evidence showed a positive association between serum prolactin and blood pressure. High serum prolactin levels were found to be associated with lower risk of type 2 diabetes in women but not in men, but evidence for an exact correlation between serum prolactin and type 2 diabetes was insufficient.

**Conclusion:** Evidence for associations between serum prolactin and metabolic profiles were insufficient. Higher serum prolactin levels might be associated with lower risk of type 2 diabetes in women. Further high-quality prospective studies are required.

## INTRODUCTION

In recent decades, metabolic disorders, including obesity, type 2 diabetes mellitus (T2DM), and atherosclerotic cardiovascular diseases have emerged as leading causes of death. The prevalence of metabolic disorders has surged since the 21^st^ century, for instance, studies showed that over one-quarter of adults were found to meet the criteria of metabolic syndrome, which is characterized as abdominal obesity, high glucose, high triglyceride (TG), low high-density lipoprotein cholesterol (HDL-C) levels and hypertension, in the last decade^[1]^. However, the pathophysiological mechanisms for the development of many metabolic disorders remained unclear. Though there is a consensus that aging and overweight contribute to most of the metabolic disorders, hormones like prolactin (PRL) could also play an important role. PRL is a polypeptide hormone given its name by stimulating lactation, and it is found to play a role in numerous biological functions including reproduction, immune response, osmoregulation, and glucose and lipid metabolism^[2, 3]^. Studies have found several mechanisms on how hyperprolactinemia interact with metabolic markers^[4]^. For instance, Dopamine-D2-receptor antagonists, like olanzapine or risperidone could promote PRL secretion and increase the risk of hyperglycemia compared to typical antipsychotics^[5]^. Nevertheless, most of the researches was conducted on patients with pathological hyperprolactinemia, like schizophrenia patients taking atypical antipsychotics^[5-7]^.

Serum PRL, in physiological range, was also found to be associated with blood pressure, blood glucose, and blood lipids recently. Studies on rodents have shown that serum PRL promotes adipose tissue function and decreases the risk of obesity and diabetes ^[8, 9]^. A study conducted in 27 normal subjects and 23 hypertension patients found that serum PRL concentration was higher in hypertension patients^[10]^. And as we mentioned above, hyperprolactinemia was found to contribute to T2DM in schizophrenia patients taking atypical antipsychotics as well via the Dopamine-D2R pathway^[5]^. However, animal studies found the effect of PRL on T2DM might be different when PRL levels were low and high, as serum PRL may play a protective role in physiological range^[4]^. A prospective cohort study enrolling 8615 women also found a negative trend for serum PRL and the risk of T2DM^[11]^.

Therefore, it is necessary to look into the association between serum PRL and metabolic disorders in physiological range. The aim of this systematic review is to review and evaluate the evidence of the association between serum PRL and metabolic profiles.

## METHODS

### 1. Literature search

PubMed, EMBASE, and the Cochrane Library were used to identify relevant studies. Two reviewers scanned all titles and abstracts identified through databases independently and make the decisions. A combination of the following terms was used for several searches: “PRL”, “prolactin”, “bodyweight index”, “waist circumference”, “obesity”, “blood pressure”, “hypertension”, “triglyceride”, “cholesterol”, “HDL”, “LDL”, “glucose”, “glycemia”, “insulin resistance”, “type 2 diabetes”, “noninsulin-dependent diabetes”, “non-insulin-dependent diabetes”, “NIDDM”, “T2DM”, “MODY”, “Maturity Onset Diabetes”, “Adult Onset Diabetes Mellitus”. References of the relevant studies were also screened.

### 2. Inclusion criteria

Include in the review were: (1) Observational studies of cross-sectional, case-control, prospective and retrospective design, and involving adults (over 18 years) were included. (2) Studies with results of odds ratios (OR) or relative risks (RR) of T2DM, or measuring at least one of the following items: waist circumference (WC), body mass index (BMI), blood pressure, high-density lipoprotein cholesterol (HDL-C), low-density lipoprotein cholesterol (LDL-C), fasting plasma glucose (FPG), glycated hemoglobin (HbA1c), homeostasis model assessment of insulin resistance (HOMA-IR). (3) Studies published in peer-reviewed journals and with an available English full-text.

### 3. Exclusion criteria

Studies with the following reasons were excluded: (1) Conference abstracts, animal studies, case reports, and reviews were excluded. (2) Studies without data of OR or correlation coefficient between serum PRL and metabolic markers were excluded. (3) Studies did not divide PRL into quartiles and did not provide OR or RR of T2DM in each quartile were excluded. (4) Studies did not provide certain serum PRL values or the value exceeded the reference range (25.0 ng/mL)^[12]^ were excluded.

### 4. Data extraction and quality assessment

The data extracted from each article included year of publication, study design, details of study participants, and main outcomes. Details of study participants included sample size, year of follow-up, number of each group, mean age and sex, mean or median PRL concentration. Main outcomes included ORs or RRs of T2DM, or correlation coefficient or ORs between serum PRL and metabolic markers (BMI, WC, blood pressure, concentration of LDL-C, HDL-C, HbA1c, FPG, HOMA-IR). Qualities of case-control and cohort studies were assessed using the Newcastle-Ottawa Scale (NOS)^[13]^, a study over 5 stars would be considered as high quality, and cross-sectional studies were assessed via Agency for Healthcare Research and Quality (AHRQ) standard^[14]^.

## RESULTS

Fig. 1 shows the flowchart of the search result. A total of 340 records were identified through database searching and 2 records through other sources. After removal of duplicates, the number was 218. Next, 191 records were excluded as they were irrelevant to our review after screening the title and abstract of each article. Full texts of the remaining studies were obtained for eligibility assessment. As a result, 14 studies were included in this review^[11, 15-27]^.

**Figure 1.**
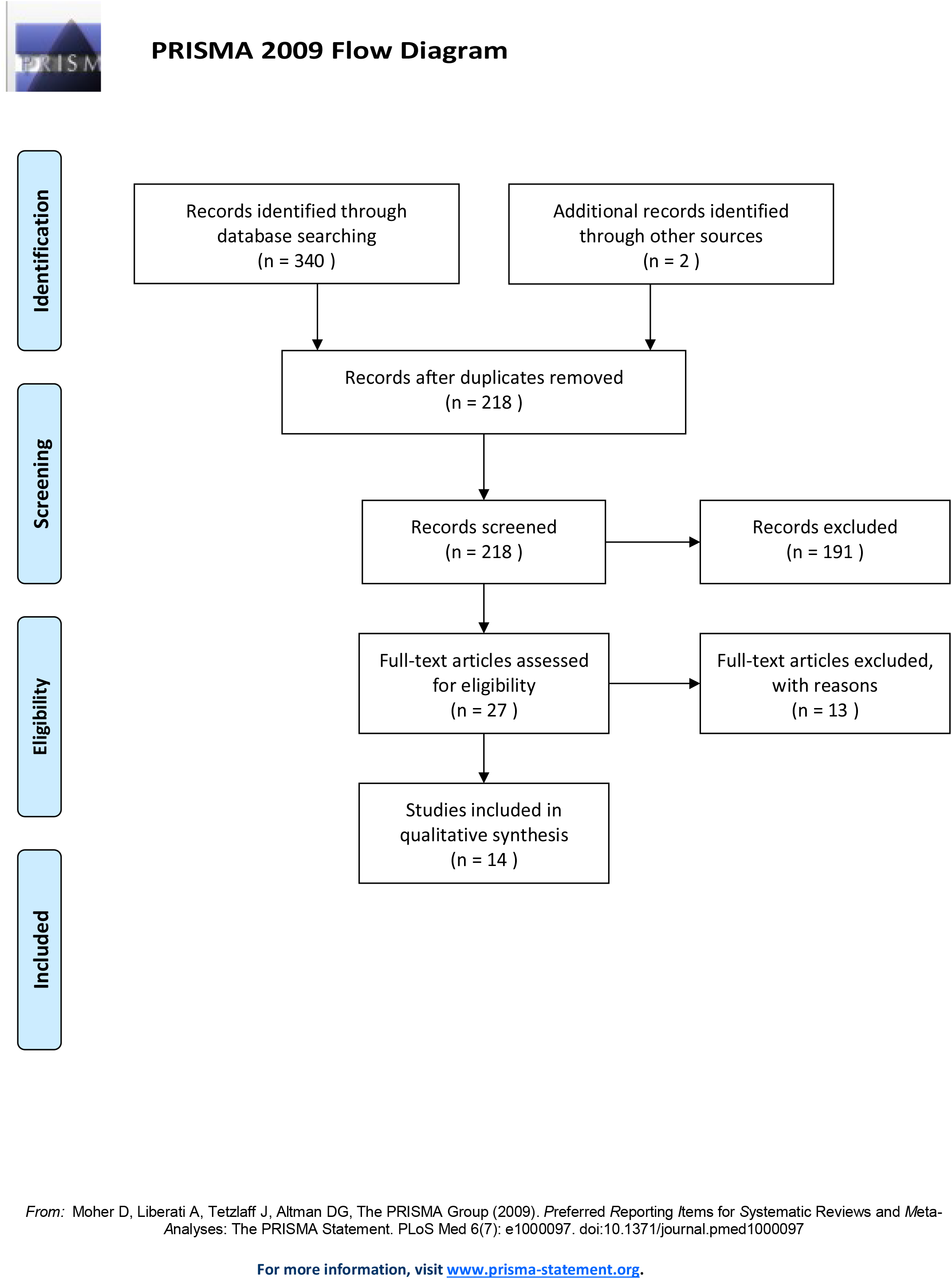

### 1. Association between serum PRL levels and metabolic markers

Nine studies were included with results on associations between serum PRL levels and BMI, WC, blood pressure, blood lipids, blood glucose, and insulin resistance ^[15, 18-24, 27]^. Among these studies, there were 3 prospective studies, 2 case-control studies, and 3 cross-sectional studies. The characteristics of studies on serum PRL and metabolic markers are summarized in Table 1. The main outcomes of each study are presented in Table 2.

**Table 1.** Characteristics of studies evaluating the association serum PRL and metabolic markers.

| Study ID | Study design | Follow up | Sample size | Participants, Gender | Age of participants (years) | NOS or AHRQ score |
| --- | --- | --- | --- | --- | --- | --- |
| Ernst. (2009) | Cross-sectional | N/A | 344 | Patients after bariatric surgery, W: 234; M: 110 | W: 40.7±11.1; M: 41.7±12.1. | 8/11 |
| Therkelsen. (2016) | Cohort | 6.1 (5.9-6.4) years | 3232 | W: 1685; M:1547 | W: 40.2; M: 40.7 | 7/9 |
| Glintborg. (2014) | Case-control | N/A | 1123 | Patients with PCOS, W: 1123 | P: 30 (23-36); C: 28 (24-47) | 5/9 |
| Ponce. (2020) | Case-control | N/A | 40 | Patients after abdominal surgery, W: 20; M: 20 | L: 42±17; H: 35±10 | 6/9 |
| Albu. (2016) | Cross-sectional | N/A | 322 | Patients with POCS, W: 322 | 24(17-31) | 7/11 |
| Zhang. (2010) | Cohort | 8 years | 874 | Postmenopausal women, W: 874 | L: 60(54-64); H: 58(54-63) | 6/9 |
| Georgiopoulos. (2017) | Cohort | 25 months | 201 | Postmenopausal women, W: 201 | 56.1(50.4-61.8) | 6/9 |
| Wagner. (2014) | Cross-sectional | N/A | 1683 | W: 1121; M: 562 | W: 38(29-50); M: 40(29-51). | 9/1 |
| Daimon. (2017) | Cross-sectional | N/A | 370 | 52.0±14.8 | M: 370 | 9 |

PRL=prolactin; W=woman; M=man; P=patients; C=control; L=low PRL concentration; H=high PRL concentration; N/A=not applicable; PCOS=polycystic ovarian syndrome.
Data for continuous variables are shown as mean ± SD or median (interquartile range).

**Table 2.** Main outcomes of studies evaluating the association serum PRL and metabolic markers.

| Study ID | PRL concentration | Main outcome |
| --- | --- | --- |
| Ernst. (2009) | W: 9.0±4.8; M: 7.9±3.6 ng/mL. | No association between BMI and PRL ($r=-0.03$, $p=0.64$), TG, TC, LDL-C, HDL-C, TC/HDL-C ratio (all $r<0.03$, $p>0.20$), fasting glucose ($r<0.03$, $p>0.20$). No association between PRL and HOMA-IR($r<0.03$, $p>0.20$). |
| Therkelsen. (2016) | W: 11.9±5.2; M: 8.2±2.9 ng/mL. | No association between BMI and PRL ($p\geq0.13$). A 5 mg/dl increment in PRL in men was associated with higher odds of hypertension (OR 1.36, 95%CI 1.08-1.71, $p<0.01$) and low HDL-C in women (OR 1.50, 95%CI 1.18-1.91, $p=0.001$). |
| Glintborg. (2014) | P: 7(5-10); C: 9 (7-13) ng/mL. | PRL in patients of PCOS was inversely associated with WC ($r=-0.13$, $p<0.05$), LDL ($r=-0.15$, $p<0.05$), TG($r=-0.14$, $p<0.05$), and positive with HDL($r=0.11$, $p<0.05$). |
| Ponce. (2020) | L: 7.4±2.3; H: 24±12 ng/mL. | PRL are not associated with WC ($r=-0.002$, $p=0.990$), SBP ($r=-0.179$, $p=0.352$), DBP ($r=-0.029$, $p=0.883$), TG ($r=-0.083$, $p=0.637$), HDL ($r=-1.132$, $p=0.512$), fasting glucose ($r<-0.110$, $p=0.5$). PRL are negatively associated HOMA-IR ($r=-0.30$, $p=0.05$) |
| Albu. (2016) | 12.9 (4.65-21.15) ng/mL. | PRL was inversely associated with HOMA-IR ($r=-0.30$, $p=0.05$), BMI ($r=-0.194$, $p<0.0001$); WC ($r=-0.217$, $p<0.0001$), and fasting glucose ($r=-0.123$, $p=0.037$). |
| Zhang. (2010) | 8.7(6.7-11.4) ng/mL. | PRL was positively associated with risk of hypertension, RR of a 1-SD higher PRL was 1.31 (95% CI 1.05-1.62, $p=0.01$). |
| Georgiopoulos. (2017) | 8.1(6.6-10.4) ng/mL. | Baseline PRL levels increased by 23.5% (OR=1.235, $p=0.001$) the odds of new-onset hypertension. |
| Wagner. (2014) | M: 8.0(6.0–12.0); W: 7.0 (5.0-9.0) ng/mL. | PRL was negatively associated with 2h-OGTT ($r=-0.26$, $p<0.0001$) and HbA1c ($r=-0.12$, $p<0.0001$). |
| Daimon. (2017) | 7.06±3.37 ng/mL. | PRL was positively associated with HOMA-IR under 12.4 ng/mL ($r=0.119$, $p<0.05$). |

PRL=prolactin; SD=standard deviation; RR=relative risk; OR=odds ratio; PCOS=polycystic ovarian syndrome; BMI=body mass index; WC=waist circumference; TG=triglyceride; TC=total cholesterol; LDL-C=low-density lipoprotein cholesterol; HDL-C=high-density lipoprotein cholesterol; SBP=systolic blood pressure; DBP=diastolic blood pressure; OGTT=oral glucose tolerance test; HbA1c=glycated hemoglobin; HOMA-IR=homeostasis model assessment of insulin resistance.
Data for continuous variables are shown as mean ± SD or median (interquartile range).

Five studies investigated the association between serum PRL and BMI, WC^[15, 19, 21-23]^. There was a significant inverse association between serum PRL and BMI, WC in 2 studies^[15, 21]^. Glintborg et al. reported that serum PRL was inversely associated with WC (*r*=-0.13, *P*<0.05) in patients with polycystic ovarian syndrome (PCOS)^[21]^. Albu et al. showed that serum PRL was negatively associated with BMI (*r*=-0.194, *P*<0.0001) and WC (*r*=-0.217, *P*<0.0001)^[15]^. However, the remaining 3 studies showed no associations between serum PRL and both BMI and WC^[19, 22, 23]^.

Four studies investigated the association between serum PRL and blood pressure^[20, 22, 23, 27]^. Of these studies, 3 reported that serum PRL increased the OR of hypertension^[20, 23, 27]^, while Ponce et al. found no associations between serum PRL with systolic blood pressure (SBP) (*r*=-0.179, *P*=0.352) and diastolic blood pressure (DBP) (*r*=-0.029, *P*=0.883)^[22]^.

Four studies investigated the association between serum PRL and blood lipids^[19, 21-23]^. Two studies found that serum PRL was negatively associated with LDL, TG, and positively associated with HDL^[21, 23]^. Therkelsen et al. found that serum PRL was positively associated with the change in total cholesterol (TC) (*r*=0.05, *P*<0.05) in women after follow-up, and a 5-mg/dL increment in serum PRL was associated with higher odds of low HDL cholesterol in women (OR=1.50, 95% confidence interval 1.18–1.91, *P*=0.001). However, there was no association between serum PRL and change in HDL-C, TG in both men and women, and TC in men^[23]^. Glintborg et al. reported that serum PRL was inversely associated with TC (*r*=-0.13, *P*<0.05), TG (*r*=-0.14, *P*<0.05), LDL (*r*=-0.15, *p*<0.05), and positively associated with HDL (*r*=0.11, P<0.05)^[21]^ in patients with PCOS. The remaining 2 studies showed that serum PRL was not associated with TC, TG, HDL, and LDL^[19, 22]^.

Four studies investigated the association between serum PRL and blood glucose^[15, 19, 22, 24]^ and two studies found that serum PRL was inversely associated with glucose metabolism^[15, 24]^. Albu et al. found serum PRL was inversely associated with fasting glucose (*r*=-0.123, *P*=0.037) in patients with normal glucose metabolism, but not in patients with T2DM, impaired fasting glucose (IFG) and impaired glucose tolerance (IGT)^[15]^. Wagner et al. found that serum PRL was negatively associated with area under curve of glucose (AUC) during the 2h-oral glucose tolerance test (2h-OGTT) (*r*=-0.26, *P*<0.0001) and HbA1c (*r*=-0.12, *P*<0.0001)^[24]^. The remaining 2 studies showed no such association in serum PRL and glucose^[19, 22]^.

Four studies reported the association between HOMA-IR and serum PRL^[15, 18, 19, 22]^. Of the presented studies, Ponce et al. (*r*=-0.30, *P*=0.05) and Albu et al. (*r*=-0.185, *P*=0.002) found a negative association between serum PRL and HOMA-IR^[15, 22]^ while Ernst at al. found no such association (*r*<0.03, *P*>0.20)^[19]^. And Daimon et al. presented a non-linear regression model as serum PRL was positively associated with HOMA-IR under 12.4 ng/mL (*r*=0.119, *P*<0.05) and such association became negative over 12.4 ng/mL (*r*=-0.199, *P*>0.05) ^[18]^.

### 2. Serum PRL levels and risk of T2DM

Five studies reported the association between T2DM and serum PRL^[11, 16, 17, 25, 26]^. The characteristics of the studies involving risk of T2DM and serum PRL are summarized in Table 2. In Table 3 a summary of outcome and results from each study are presented. Four out of the 5 studies showed that higher serum PRL coincided with a lower risk of T2DM in women^[11, 17, 25, 26]^, but the outcomes were inconsistent in men. Wang et al. showed an inverse association in men in their cross-sectional study in 2013^[25]^ but a positive association in their prospective study in 2016^[26]^. Chahar et al. also gave a negative association in their study^[17]^, while Balbach et al. showed a positive association^[16]^. Moreover, Balbach et al. reported the OR of T2DM with per standard deviation (SD) increment of serum PRL (OR=1.13, 95% confidence interval 0.89-1.44, *P*>0.05)^[16]^.

**Table 3.** Characteristics of studies on serum PRL and type 2 diabetes.

| Study ID | Wang.<br>(2016) | Chahar.<br>(2017) | Balbach.<br>(2013) | Li.<br>(2018) | Wang.<br>(2013) |
| --- | --- | --- | --- | --- | --- |
| Study design | Cohort | Cross-sectional | Cohort | Cohort | Cross-sectional |
| Follow up | 3.7 yrs | N/A | 5.0 yrs | 22 yrs | N/A |
| Sample size | 2377 | 300 | 3993 | 8615 | 2377 |
| Participants,<br>Gender | W: 1343;<br>M; 1034. | W: 120 ;<br>M: 180 | W: 2027;<br>M: 1966 | Nurse,<br>W: 8615 | W: 1343;<br>M: 1034 |
| NOS or<br>AHRQ score | 7/9 | 8/11 | 6/9 | 8/9 | 9/11 |

PRL=prolactin, N/A=not applicable, W=women, M=men

**Table 4.**
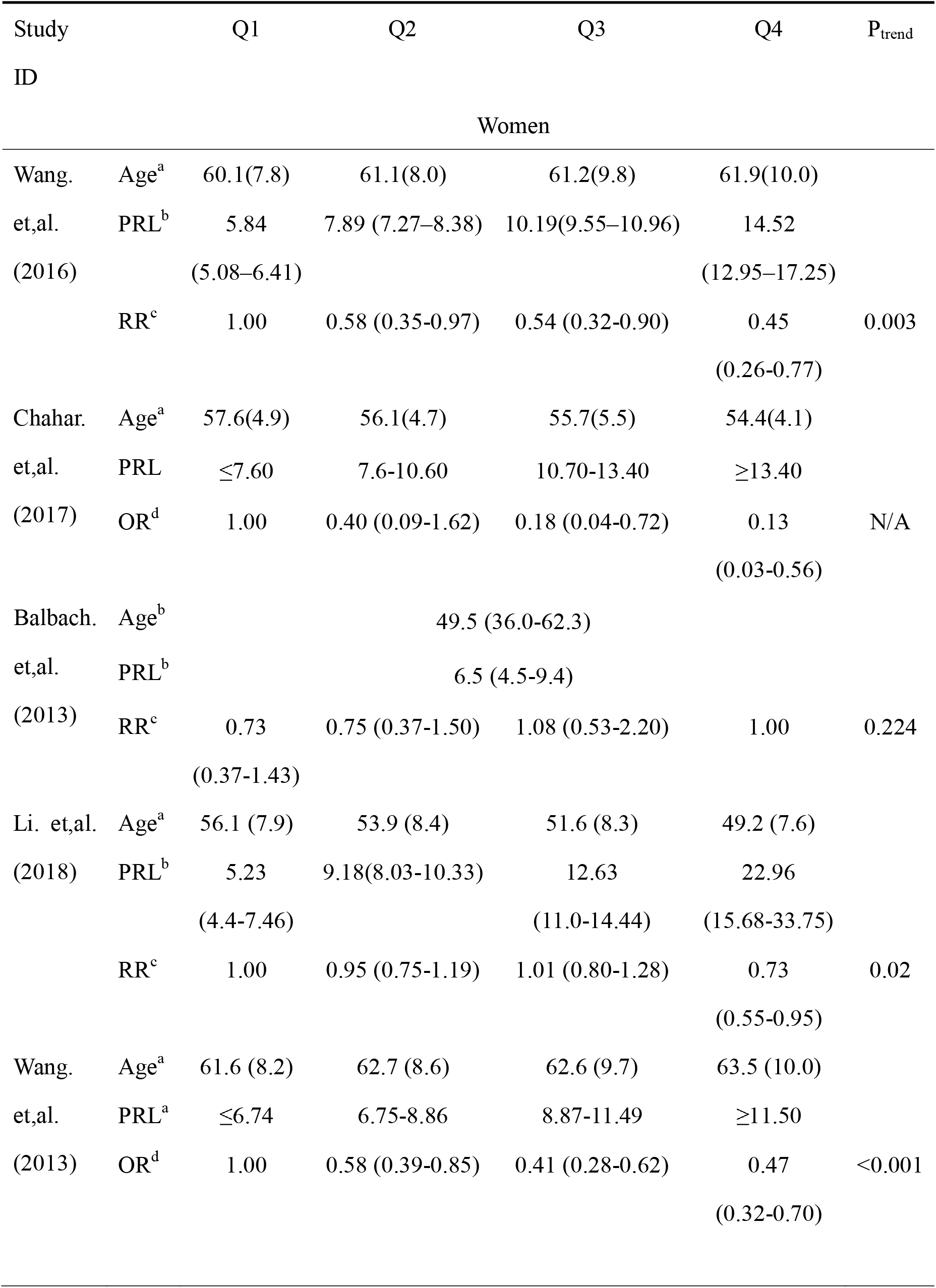
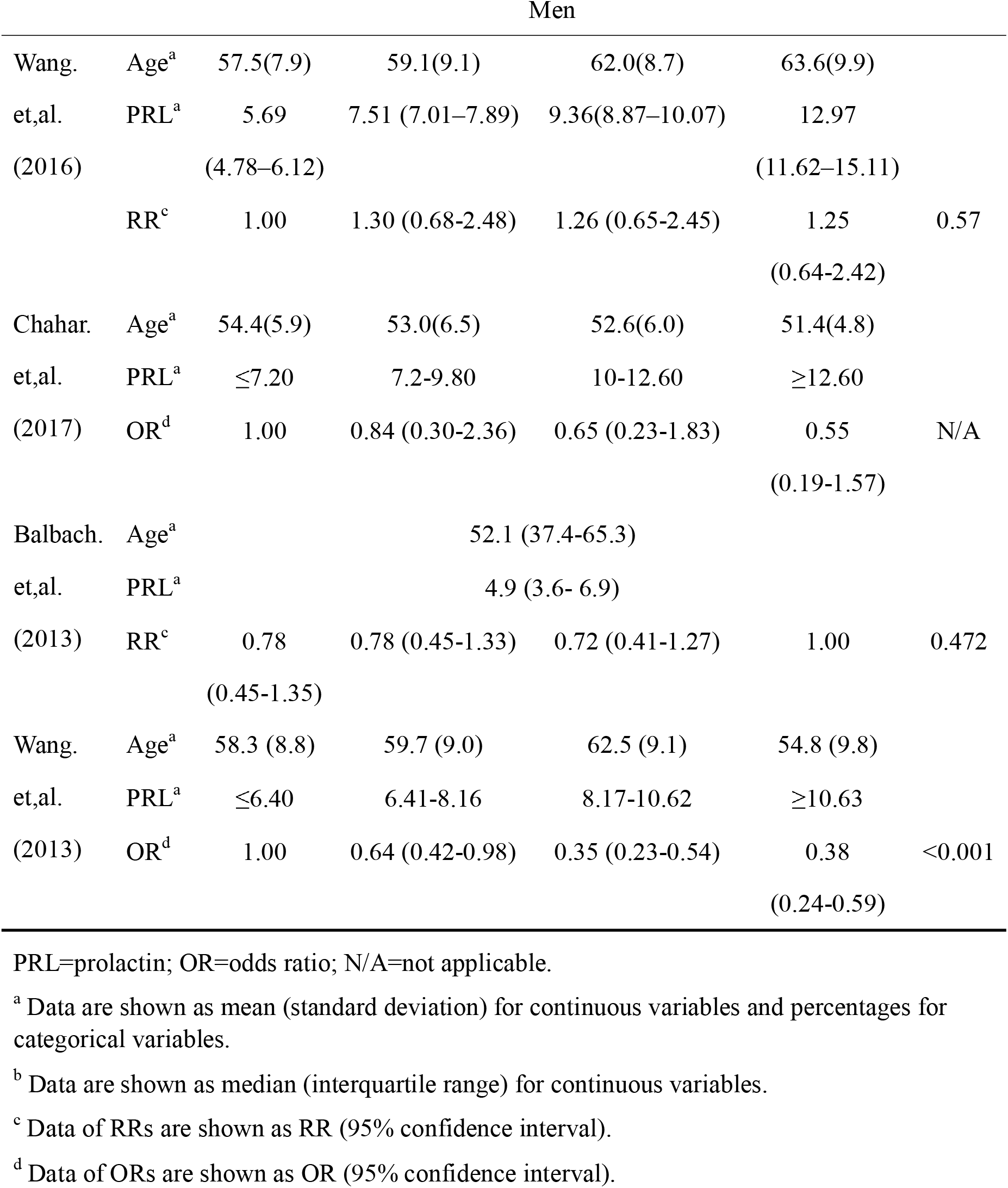
Main outcomes of each study on serum PRL and type 2 diabetes. PRL=prolactin; OR=odds ratio; N/A=not applicable.

## DISCUSSION

Studies on rodents have shown an inverse association in serum PRL and insulin resistance, obesity, and diabetes. Different mechanisms have been supposed: Serum PRL decreases lipogenesis, reduces adipocyte hypertrophy, increases adipocyte hyperplasia, enhance adipocyte function^[9, 28]^; promotes proliferation, survival, and insulin secretion of pancreatic β-cells^[29]^; decreases glucose uptake by inhibiting glucose transporter 4(GLUT4) mRNA expression^[30]^. These mechanisms may ultimately contribute to metabolic disorders including obesity and T2DM in animals with low serum PRL levels.

However, in this systematic review of serum PRL and metabolic disorders, there was only moderate evidence on serum PRL and T2DM supported by 2 high-quality prospective studies that higher serum PRL levels were associated with a lower risk of T2DM in women. Evidence for hypertension and serum PRL could also be considered as moderate. Results on other metabolic markers were limited due to both the number and quality of the included studies.

Five studies investigated the association between T2DM and serum PRL and all of these studies showed a negative trend for higher serum PRL levels and lower risk of T2DM in women^[11, 16, 17, 25, 26]^. A total of 15285 individuals were enrolled in these studies, and the maximum follow-up period was 22 years. Populations from four studies were community originated^[16, 17, 25, 26]^ while the remaining one were nurses^[11]^. Besides, two prospective studies had their response rate over 95%^[11, 26]^. However, such association might be non-linear, as Balbach et al. reported that data about OR of T2DM with per SD increment in serum PRL^[16]^ was not significant. Similarly, among the included studies, although the difference of OR between the fourth quartile and first quartile was significant, Wang et al. found it become less significant compared to the third quartile^[25]^. Similar results were reported by Li et al. as OR of the third quartile was found insignificant compared to the first quartile^[11]^. Besides, we noticed that the mean concentration of serum PRL of the fourth quartiles from Li et al.^[11]^ was over 22 ng/mL, a value that almost reaches the reference range of serum PRL^[12]^, which might indicate a different effect of serum PRL in women with high serum PRL levels.

The evidence for an association in serum PRL and T2DM in men remained insufficient, as only Wang et al. reported a significant trend in serum PRL and T2DM in their study^[25]^. Their study found that serum PRL was negatively associated with T2DM in men in the cross-sectional design. Nevertheless, the remaining 3 studies did not show a significant trend for serum PRL and T2DM in men, and prospective studies from Wang et al. and Balbach et al. even found that OR of the fourth quartile was higher than the first quartile^[16, 26]^. Interestingly, Daimon et al. found that the association between serum PRL and HOMA-IR was concentration-dependent in men. They found a peak point of the regression curve at a serum PRL level of 12.4 ng/mL, and above this point serum PRL was less significantly associated with HOMA-IR^[18]^. Insulin resistance is an important aspect of T2DM, therefore, their study may indicate a different role of serum PRL in T2DM in men.

Data about OR of T2DM with per SD increment in serum PRL was not presented in 4 studies^[11, 17, 25, 26]^, which allowed an additional quantitative synthesis. Furthermore, the effect of serum PRL on T2DM might be complex within the normal range as we mentioned above. Therefore, studies with stratified analysis based on serum PRL levels and data of OR with per SD increment in serum PRL were also required.

In this review, the evidence for an association between serum PRL and BMI or WC, TG, LDL, or HDL, blood glucose was insufficient. Five studies investigated the association between BMI, WC, and serum PRL, and 3 of them reported no significant association^[19, 22, 23]^. Meanwhile, both of the remaining 2 studies were conducted in PCOS patients, demonstrating a negative association between serum PRL and BMI or WC ^[15, 21]^. And there are only 4 studies investigated LDL and TG, with 2 studies finding no significant association between LDL, TG, and serum PRL^[19, 22]^. Similar results were found in serum PRL and blood glucose.

It could be the cases that there are several limitations in this review. There are only 3 prospective studies investigated metabolic markers and serum PRL, while 2 of which only investigated the association in blood pressure and serum PRL^[20, 27]^. All of the 3 studies found serum PRL was positively associated with the incidence of hypertension^[20, 23, 27]^

And we did not exclude studies conducted on PCOS patients, postmenopausal women, or obese patients after surgery, due to the sample size of the included studies, on the one hand. On the other hand, the values of PRL concentration in these studies are within the reference range we mentioned above, while the values in patients with hyperprolactinemia exceed the upper limit^[6, 7]^. It is reported that hyperprolactinemia contributes to T2DM morbidity, thus we hypothesize that the effect of serum PRL in physiological range on metabolic markers is different. However, it seems that other factors in these studies, like health status and age matters. We found the results were more consistent within the 2 studies conducted on PCOS patients, compared to other studies, which might indicate a different role that PRL plays in PCOS patients. In this review stratified analysis is not applicable limited by the quantity of the included study, therefore, more studies on healthy populations are required regarding this issue. The results from current studies are impossible to demonstrate a certain association between PRL and these metabolic markers.

There would be some reporting bias as well, as only 3 databases were used for literature search, and only 2 reviewers scanned and identified the records. Only published studies with an available English full-text were included. Because of the limitation in both sample size, study design, and study population of the included studies, more high-quality studies could be helpful.

## Data Availability

The data used to support the findings of this study are available from the corresponding author upon request.

## CONFLICT OF INTEREST

No potential conflict of interest relevant to this article was reported.

## Notes

### Competing Interest Statement

The authors have declared no competing interest.

### Funding Statement

None.

### Author Declarations

Not applicable. Systematic review.

